# HABITAT: Evolution of a Cloud-Based, Mesh-Enabled Research Data Ecosystem at an Academic Medical Center

**DOI:** 10.64898/2026.07.01.26357043

**Authors:** Yaswitha Jampani, Kamruz Rana, Saber Hossain, Vasanthi Mandhadi, Soliman Islam, Tamara M. McMahon, Xiaofan Niu, Song Xing, James McClay

**Affiliations:** University of Missouri, Columbia, Missouri, USA; University of Missouri, Kansas City, Missouri, USA

## Abstract

Academic medical centers face growing demand for clinical data access that traditional informatics infrastructure cannot support at scale. We describe HABITAT, a governed, tiered research data ecosystem at the University of Missouri integrating internal and external sources, including unstructured clinical data, into a layered architecture supporting PCORnet and OMOP common data models. A three-tier model provides self-service feasibility tools, secure cloud analytics, and curated honest broker services under a Five Safes governance framework. From mid-2022 through February 2026, HABITAT received 1,150 requests with 689 fulfilled and supported 29 externally funded projects. Median fulfillment for internal data requests improved from 30 to 20 days, and an early survey captured 54 scholarly outputs, including peer-reviewed publications in high-impact journals. HABITAT supported coursework to multi-site research without proportional staff increases, while cost recovery and data standardization barriers highlight challenges institutions face as AI-driven data demand grows.

## Introduction

In modern healthcare research, Electronic Health Record (EHR) data serves as a backbone supporting hypothesis generation, patient recruitment for clinical trials, and large-scale analytics^.1^ At the University of Missouri (MU), as at many other institutions, investigators rely on manual data pulls from EHR systems coordinated through data analysts. Though this model is effective for isolated projects, it is labor-intensive, hard to scale, and likely to compromise reproducibility. Over time, self-service tools such as Informatics for Integrating Biology and the Bedside (i2b2)^2^ have been introduced at MU to support feasibility assessments and cohort discovery. These platforms use structured ontologies and query-based interfaces, making data easily accessible. However, ontology limitations, restrictive data domains, and performance constraints made them poorly suited for end-to-end research studies that require more granularity.

Meanwhile, MU joined national networks such as the Greater Plains Collaborative (GPC)^3^, a National Patient-Centered Clinical Research Network (PCORnet^®)4^ affiliate and Evolving Network for Encapsulating Clinical and Translational Research (ENACT)^5^, expanding capacity for collaborative research. To overcome the limitations of ad hoc data pulls and i2b2, the Center for Biomedical Informatics (CBMI) at MU School of Medicine adopted an Enterprise Data Warehouse for Research (EDW4R) Framework.^6,7^ The system was built on EHR data, using the standard Common Data Models (CDM) as its foundational structure. This transition was an initiative to help data analysts and researchers with governed and standardized clinical data, benefitting both local and collaborative research. Nevertheless, this standardization of CDM can strip out the EHR nuances and data elements that don’t map cleanly to the model, compromising the granularity of the original data.^8^ As research grew, we observed the need to include unstructured clinical text, genomics-derived data, and external socioeconomic variables, which traditional models alone could not support. Growing use of large language models (LLMs) and big data analytics introduced technical and computational demands,^9^ prompting CBMI to build a unified HIPAA-compliant cloud ecosystem. This led to the development of Health Analytics and Biomedical Informatics Trusted Access and Technology (HABITAT).

HABITAT is a single governed research ecosystem with extensions to support end-to-end investigative pipelines for hypothesis generation and grant development through large-scale analytics and LLM-based applications. HABITAT delivers research support through a three-tiered service model from self-service feasibility tools to secure cloud analytics and curated data services based on individual project needs. This paper describes the HABITAT architecture, data workflows, governance model, and early impact of its research services.

### HABITAT System Architecture

HABITAT is covered under an approved MU IRB detailing data sources, team members, and standard operating procedures (SOP). The SOP covers the process for the creation, maintenance, enhancement, and sharing of data and application resources for supporting educational and research activities. Data types and sources are defined as well as the date shifting algorithm used during the de-identification process.

### HABITAT Data Warehouse and Data Lake

HABITAT operates a hybrid cloud research data infrastructure that distributes workloads across Amazon Web Services (AWS) and Snowflake. AWS functions as the foundational layer for staging and sharing large-scale datasets, hosting applications, and executing computationally intensive workloads. To accommodate varying pipeline demands, this AWS layer utilizes a flexible combination of managed services, containerized environments, and event-driven compute. Snowflake then serves as the primary engine for downstream data transformation, integration, and analysis. This design uses Snowflake’s ability to separate storage and compute, scale independently of data volume, and handle concurrent workloads. It ensures controlled data sharing across isolated environments, secured by strict role-based access and comprehensive audit logging.

### Bronze Layer (Source Data)

Multiple heterogeneous source systems feed HABITAT, including electronic health records, billing, pharmacy, tumor registry, health information exchange, genomics, socioeconomic and environmental variables,^10^ and national death master files. Data from all sources are ingested in their original form with minimal transformation. Each source type is processed through a standardized pipeline designed for its specific structure and format, ensuring consistency across different systems before being combined in later stages. The layer refreshes nightly to weekly through incremental loads with automated pipelines pushing updates to all dependent layers.

### Silver Layer (HABITAT Data Model)

From the bronze layer, source tables are integrated into the HABITAT data model (**Figure 1**), where patient linkages are established across systems using a master patient index. This layer is standardized based on CDMs such as the PCORnet^4,11^ and OMOP^12^ as its foundational structure for applicable domains and tables. Data elements outside the standard CDM scope are designed with additional standardization. In this layer, data undergoes quality assessment, vetting, and logical curation to produce a research-ready data layer. The HABITAT data model provides standardization of real-world medication, laboratory, clinical note, and other clinical observations data into standard terminology from RxNorm, LOINC, SNOMED, LOINC document ontology,^13^ and other standard ontologies. This produces a unified patient-centered data model that eliminates the need to query source systems separately, supporting research across all integrated data sources.

**Figure 1.**
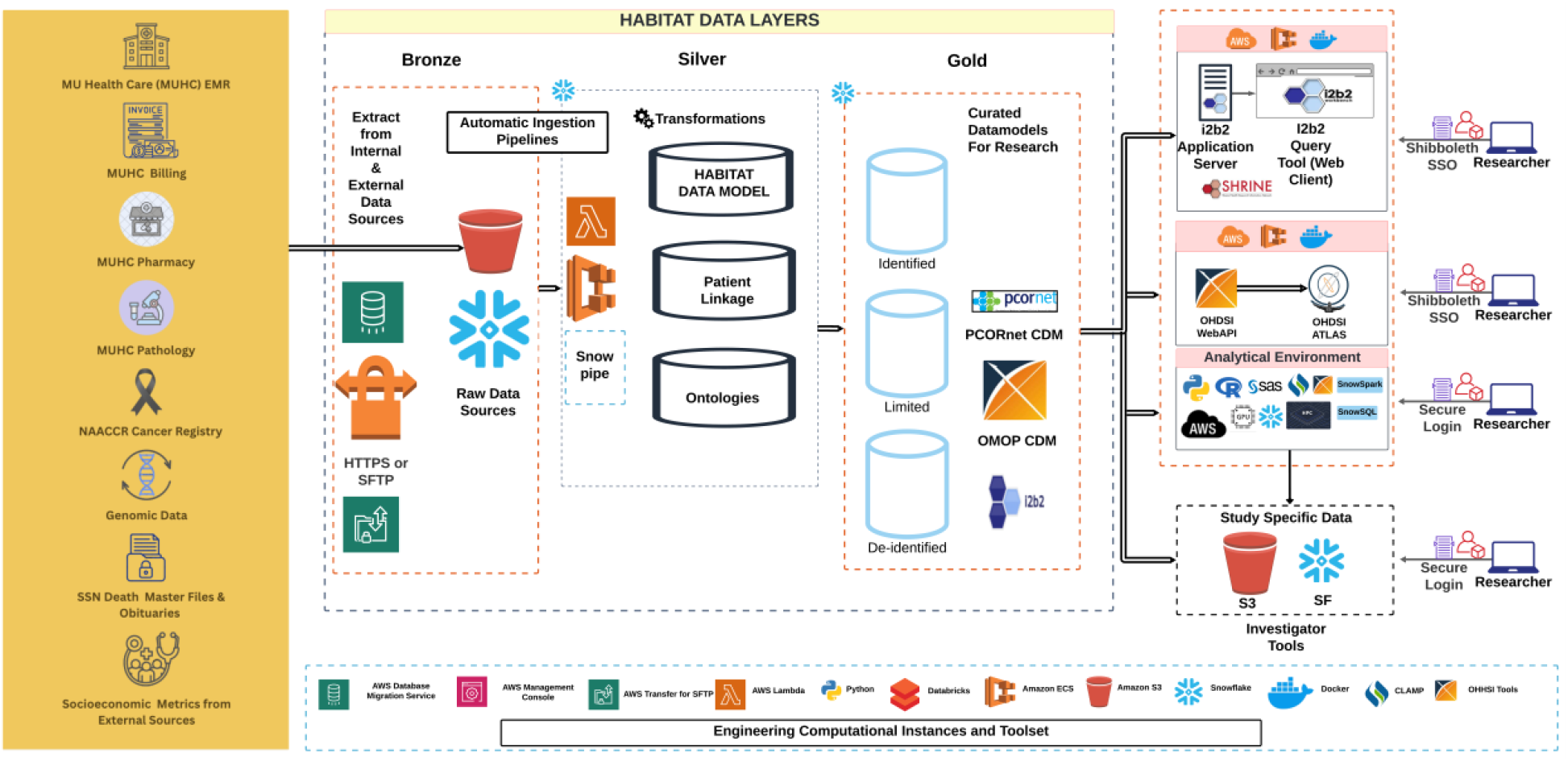
HABITAT Research Data Warehouse Architecture. Data flows from source systems through extraction, Snowflake-based staging, and transformation into the HABITAT Research Data Warehouse, organized into identified, limited, and de-identified buckets.

### Gold Layer (Curated Data)

Based on HABITAT’s standard protocols, a retraceable gold layer is created from the silver layer, organized into three buckets. The identified bucket retains all 18 HIPAA identifiers and supports research requiring full protected health information (PHI) under appropriate IRB authorization. The limited dataset bucket applies HIPAA limited dataset standards and serves as the foundation for PCORnet and GPC^3^ participation with mappings aligned to PCORnet CDM specifications. The de-identified bucket removes HIPAA identifiers and applies patient-level date-shifting up to 365 days. All three buckets share the same foundational structure, allowing cohorts to move across buckets without rebuilding data processing steps.

### Analytical Applications

HABITAT extends the gold layer to support i2b2^2^, SHRINE^5^, ATLAS (Observational Health Data Sciences and Informatics, OHDSI), and Research Analytics & Compute Hub (RACH), with i2b2 star schema and Observational Medical Outcomes Partnership (OMOP) CDM structures populated directly from the gold layer on paired Snowflake and PostgreSQL databases^12,14^. End-user applications are hosted in an isolated AWS account without direct internet access, with ingress managed through an Application Load Balancer (ALB) and Web Application Firewall (WAF) integration.

### Unstructured Data Processing

Clinical notes are extracted from the EHR, staged in AWS S3, decompressed and transformed into queryable tables in Snowflake, then organized with patient, encounter, note type, and timestamp linkages — enabling research that integrates structured and unstructured clinical data and supporting downstream natural language processing (NLP) workflows.

### LLM and Machine Learning Pipeline

An isolated AWS-based pipeline supports LLM and machine learning workflows on clinical text within a secure environment that has no external internet access. The system operates inside a dedicated virtual private cloud, where processing instances interact only with internal AWS resources. Encrypted Amazon S3 buckets serve as the source for clinical notes, intermediate artifacts, and approved model files, while application packages and containers are delivered through AWS-native repositories and deployment services. Within this enclave, clinical notes undergo natural language processing tasks such as concept extraction, classification, summarization, and automated de-identification. Identifiable data remain confined to the secure environment, and only approved anonymized outputs are released under institutional governance. Machine learning and LLM models are staged internally, validated using synthetic datasets, and reviewed by institutional information security teams before deployment to ensure compliance with organizational and regulatory requirements.

### Cloud Infrastructure, Isolation and Security

AWS environments are continuously monitored through GuardDuty and Security Hub to detect anomalous activity, misconfigurations, and potential security threats. Monthly maintenance covers Lambda updates, pipeline reviews, vulnerability remediation, and workspace patching. New pipelines follow AWS Well-Architected Framework best practices and campus information security review. Snowflake and AWS provide additional governance through role-based access control, data masking, row-level security, time travel, and Trust Center monitoring. HABITAT’s security controls are aligned with NIST SP 800-53 moderate control standards across both AWS and Snowflake platforms.

### HABITAT Service Model

The HABITAT service model is designed around a governance-first framework, structuring data access through three tiers to serve researchers while meeting institutional compliance requirements. HABITAT’s tiered access model enables traceability across all services by integrating governance controls directly into provisioning workflows and system architecture. All tiers draw from the same underlying governed data infrastructure, maintaining data provenance throughout the research lifecycle.

#### Tier 1-Self-Service Hypothesis Generation and Feasibility

For initial feasibility studies and cohort exploration, HABITAT provides access to i2b2^2^ and ATLAS.^12^ These tools enable researchers to query patient populations, calculate incidence and prevalence, and perform cohort characterization independently, supporting hypothesis generation and feasibility assessment for study planning and grant development.

#### Tier 2-Secure Cloud Analytics and Data Access

RACH is an integrated service combining Snowflake and AWS Research Studio (RES). RACH provides secure, project-level cloud workspaces for analyzing de-identified datasets within governed environments, configured based on project storage and compute needs including GPU access.

#### Tier 3-Curated Dataset Fulfillment and Honest Broker Services

For studies requiring specific data elements, customized phenotypes, or identified datasets, the CBMI honest broker team provides direct support.^15^ In this tier, honest brokers work directly with researchers to curate datasets in the formats required by their specific regulatory approvals and security oversight. Tier 3 services extend beyond MU investigators to support externally funded and collaborative research, including network requests submitted through PCORnet^4^ Front Door and GPC^3^ line-item data requests, and multi-site projects where CBMI serves as either a participating or lead site. These requests follow the same governed curation workflow and honest broker process as internal requests, with effort tracked as a percentage of CBMI’s institutional contribution to each project.

**Table 1** summarizes the three service tiers, their primary use cases, IRB requirements, and corresponding data layers within the HABITAT architecture. Each tier draws from the same underlying governed data infrastructure, with access scoped to the appropriate analytical bucket based on regulatory requirements and project needs.

**Table 1.** Summary of HABITAT three-tiered service model and its alignment with HABITAT architecture.

| Tier | Primary Use | IRB Requirement | Data Layer |
| --- | --- | --- | --- |
| **Tier 1** | Feasibility and cohort exploration | Not required | Gold Layer de-identified bucket (aggregate counts) |
| **Tier 2** | De-identified dataset analysis | IRB approval or Non-Human Subjects Determination | Gold Layer de-identified bucket (line-item dataset) |
| **Tier 3** | Customized phenotypes | IRB approval and applicable waivers | Harmonized and all three buckets from gold layer |

### Governance

HABITAT’s governance framework^6^ aligns with the Five Safes principles,^16^ a structured approach to data governance adopted by national data services globally. Each dimension of the Five Safes maps to existing HABITAT policies, workflows, and technical controls addressing institutional, regulatory, operational, and ethical requirements (**Table 2**).

**Table 2.** HABITAT’s governance alignment with Five Safes principles.

| Safe Principle | HABITAT Implementation |
| --- | --- |
| **Safe People** | Access requests are submitted through a centralized REDCap<sup>17</sup> intake system. Non-faculty users require faculty endorsement before access is granted. CITI certification and a signed data use agreement (DUA) or service use agreement are required at intake for all users. |
| **Safe Projects** | Each request collects a project description and requires IRB approval or applicable waivers. Curated dataset requests undergo principal investigator (PI) alignment review before fulfillment proceeds. |
| **Safe Data** | Data access is scoped to tiers and requested data elements, inclusion/exclusion criteria, and standard codes are reviewed against the approved protocol before provisioning. |
| **Safe Settings** | Isolated Snowflake databases and AWS RES workspaces. The HABITAT ecosystem operates within a separate Snowflake environment, isolated from end user environments. |
| **Safe Outputs** | Prior to fulfillment, the dataset scope is verified against the approved IRB protocol. Disclosure records are maintained per service request and reconcilable by Service ID. Access expiration is tracked and audited. |

#### Centralized Intake and Triage

Requests are submitted through a centralized REDCap^**17**^ intake system, assigned within custom dashboards, and tracked across intake, review, and completion stages. When multiple services are requested, a single honest broker coordinates fulfillment to maintain consistency, auditability, and traceability.

#### Provisioning and Access Management

Tier 1 access is managed through i2b2^2^ and ATLAS^12^ administrative portals, returning obfuscated aggregate counts. Tier 2 projects are provisioned with logically isolated Snowflake databases and AWS RES workspaces named by IRB identifier, with access governed by Data Use Agreements and subject to audit logging. Researchers access standardized assets in PCORnet^4^ and OMOP^12^ formats alongside curated ontologies. Tier 3 curation occurs within a separate HABITAT Snowflake environment isolated from Tier 1 and Tier 2. For each request, a dedicated schema labeled by Service ID supports traceability, and a copy of the eligible cohort is retained within the schema where appropriate to support reproducibility. Final curated datasets are delivered through institutionally approved secure transfer mechanisms.

#### Monitoring

Access expiration is tracked through REDCap, with automated outreach prior to expiration. If no response is received within 14 business days, access is deactivated and reactivation requires a new intake request. Audit reports are reviewed monthly to verify that only users with valid approvals maintain active access.

#### Patient Disclosure Tracking

For limited or identified dataset releases, HABITAT maintains a dual-system disclosure framework to support regulatory oversight and institutional accountability. A centralized Snowflake table records patient identifiers linked to the Service ID, while detailed PHI release documentation is maintained in REDCap by the honest broker. This structure supports Accounting of Disclosures requirements while maintaining separation between patient identifiers and clinical attributes. The Service ID serves as the linking key between both systems when reconciliation is required.

### Early Impact

Since its launch in mid-2022, HABITAT has received 1,150 service requests from MU investigators across all three tiers through February 2026, with 689 fulfilled and 20 currently in progress. The remaining requests reflect researcher-initiated cancellations, withdrawals, and governance-based rejections. Of 36 rejected requests, representing 3.2% of total submissions, rejections reflect appropriate governance screening at intake rather than system failure. Request volume grew consistently from 2022 through 2025. Across all tiers, students represented the largest user role group, reflecting HABITAT’s role in supporting research programs and academic coursework. Because a single request may span multiple tiers, tier-level counts sum to more than the 1,150 total requests.

Tier 1 i2b2 received 473 requests with 330 fulfilled, with usage remaining relatively steady year over year across departments and campuses including the University of Missouri-Kansas City (UMKC), consistent with its role in self-service feasibility assessment and coursework. **Figure 2** shows query volume grew consistently from Q1 2023 through Q4 2025, with stable active users and periodic onboarding spikes in Q3 2024 consistent with course cycles.

**Figure 2.**
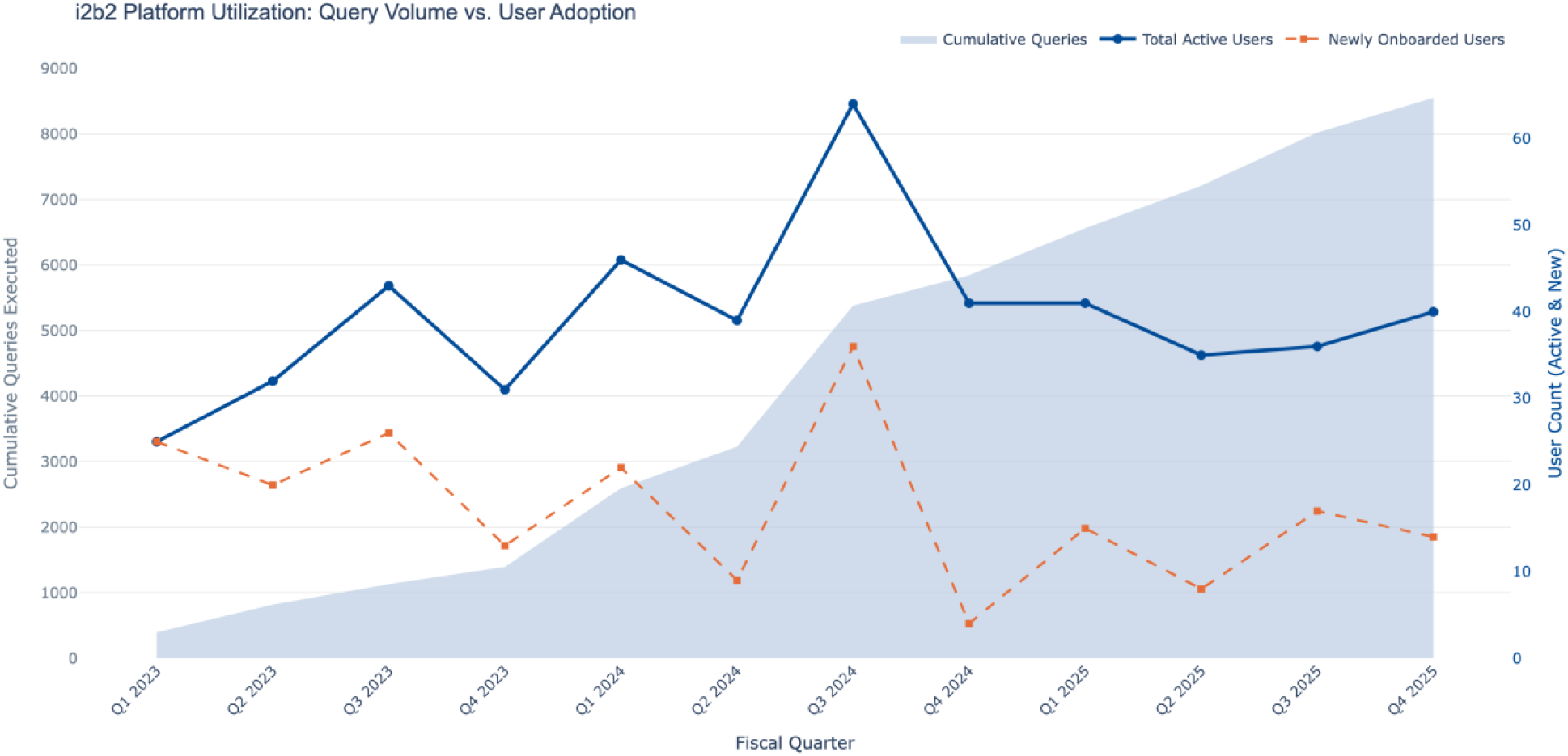
i2b2 Platform Utilization and User Adoption. A historical view comparing cumulative system query volume (shaded area, left axis) with quarterly total active and newly onboarded users (lines, right axis).

Tier 2 RACH received 327 requests with 168 fulfilled: 82 for research and 86 for departmental coursework. Graduate dissertation work was supported at no cost to researchers under CBMI’s subsidized model. Beginning in 2026, RACH moved to cost recovery to support long-term sustainability.

Tier 3 curated data services received 589 requests with 279 fulfilled from local MU investigators. Beyond local requests, Tier 3 supported 59 PCORnet Front Door and 14 GPC line-item data requests. Non-funded curated requests grew over time, suggesting more investigators are using curated services before securing formal funding. Funded local requests grew through 2023 but declined in 2024 and 2025, likely reflecting cost recovery introduction and broader reductions in federal research funding. PCORnet Front Door requests grew steadily, while GPC requests declined as a shared analytics platform reduced the need for exported datasets from participating sites. The drop in Tier 3 cancellations from 72 in 2024 to 64 in 2025 suggests clearer scope expectations at intake reduced mid-process withdrawals (**Figure 3**).

**Figure 3.**
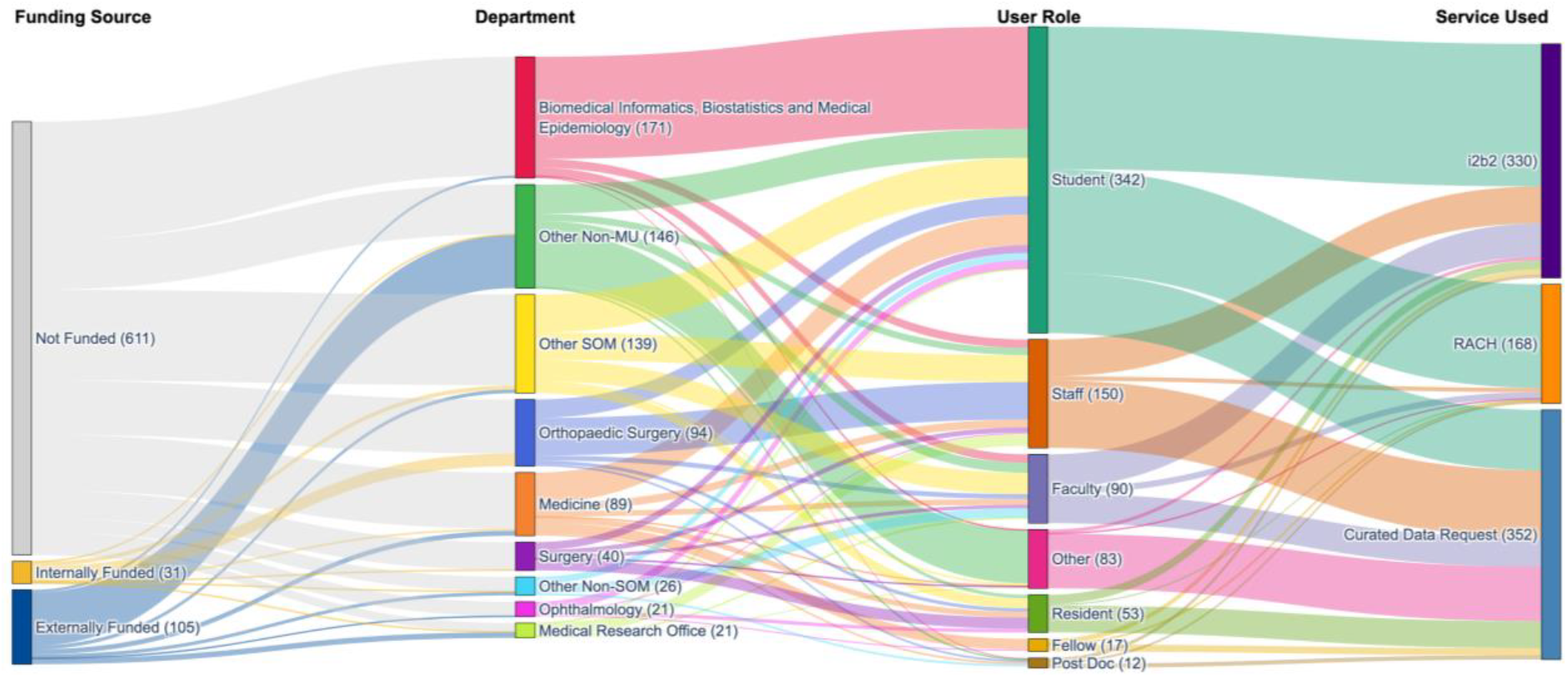
Proportional flow of completed data requests across the informatics ecosystem. The vertical height of each node and the width of each connecting band are strictly proportional to the absolute volume of completed requests. External network queries (e.g., PCORnet, GPC data requests) are integrated into the pipeline as externally funded, non-affiliated entities using mediated curated data services.

#### Service Fulfillment Times

Fulfillment times varied by tier (**Figure 4**, bottom). i2b2 requests were consistently fulfilled within approximately one week, the fastest and most predictable service. RACH provisioning remained within one to two weeks. Curated data requests had longer fulfillment times given the honest broker effort involved in phenotype curation, IRB alignment, and dataset preparation. Median fulfillment times for curated requests improved from 2022 through 2024 but increased in 2025. In 2025, funded requests were tightly clustered within approximately one month with no visible extreme outliers, whereas earlier years showed wider distributions. Requests exceeding 45 days are likely due to project complexity and governance-related delays.

**Figure 4.**
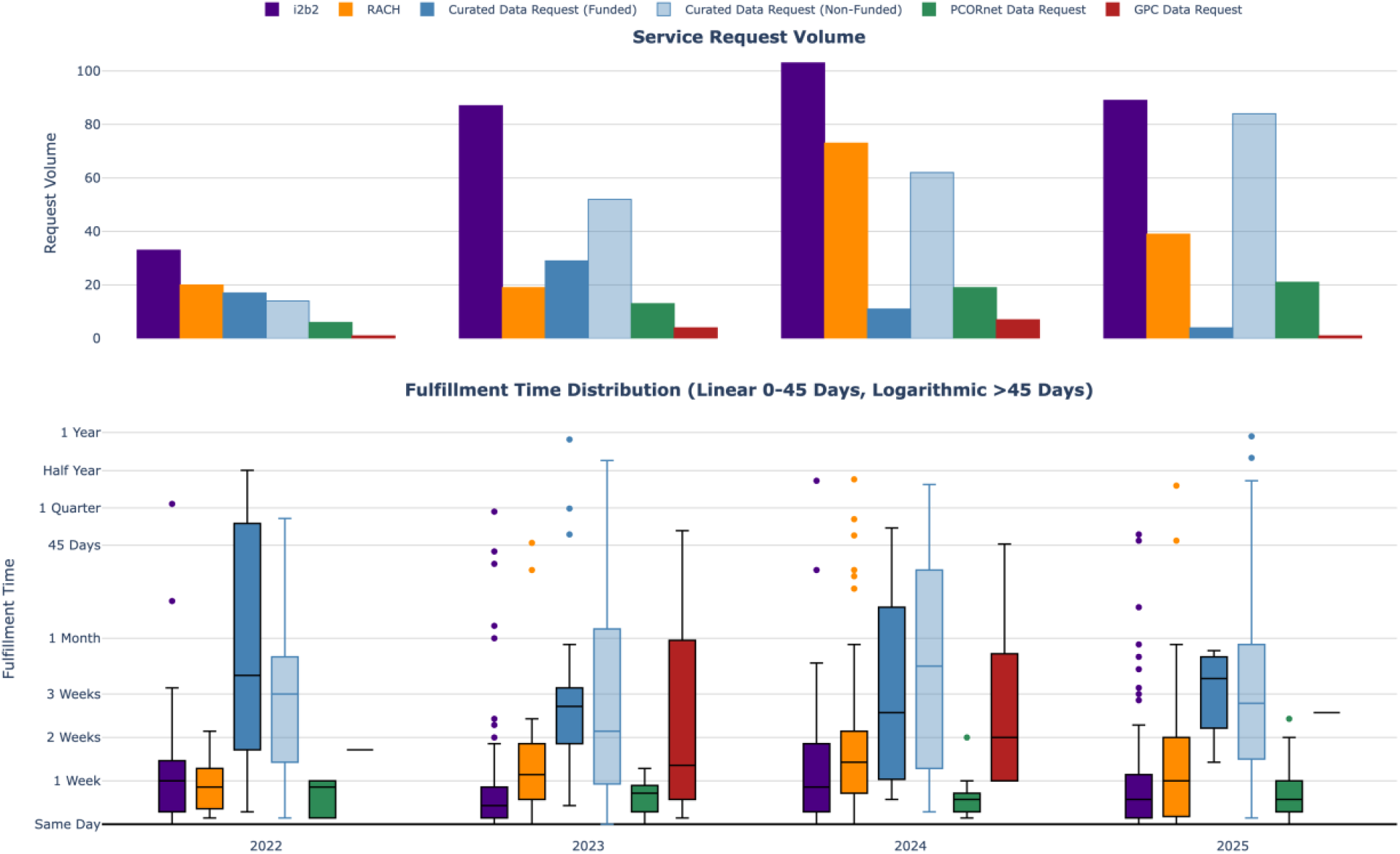
(Top) Annual request volume across self-service platforms (i2b2, RACH) and mediated data delivery pipelines (Curated, PCORnet, and GPC Data Requests). Internal mediated requests are stratified by funding status. (Bottom) Distribution of fulfillment times in days for completed requests. The y-axis utilizes a piecewise linear-logarithmic transformation (linear mapping for ≤45 days; base-10 logarithmic scaling for >45 days).

**Figure 4** presents annual service request volume by tier and funding status (top) and time of fulfillment distributions by service across years (bottom).

#### Comparison with Pre-HABITAT Workflows

To assess fulfillment efficiency relative to the pre-HABITAT era, we compared median fulfillment times for internal data requests across both periods. Prior to HABITAT (2018–2021), requests were fulfilled through nine disparate tools across three teams with no unified intake or documented turnaround standards, with a median fulfillment time of 30 days and a 90th percentile of 77 days. Following HABITAT implementation (mid 2022–February 2026), median fulfillment improved to 20 days, a 33% reduction, achieved while consolidating nine tools into a single governed pipeline and supporting a broader, more complex request mix.

#### Collaborative and Network Research Support

HABITAT also supported externally funded multi-site projects in which CBMI was a direct collaborator, serving as either a participating or lead site. All collaborative requests followed the same governed curation workflow and honest broker process as internal Tier 3 requests, with effort tracked as a percentage of CBMI’s institutional contribution to each project. From 2022 through 2025, HABITAT supported 29 unique research projects across federal and state sponsors, with approximately half (n=14) funded by NIH, nine by Patient-Centered Outcomes Research Institute (PCORI), and the remainder by Advanced Research Projects Agency for Health (ARPA-H), CDC, DoD, Missouri state agencies, and industry sponsors.

#### Scholarly Outcomes

To track research outcomes, CBMI implemented an automated publication tracking survey through REDCap Automated Survey Invitations in early 2026, sent to local service-tier researchers one year after service fulfillment. Since implementation, 77 responses have been received out of approximately 650 surveys sent (11.8%), likely undercounting actual outputs at this early stage. Self-reported scholarly activity included 54 scholarly outputs and three grant submissions, two of which were funded. HABITAT-supported local research has produced peer-reviewed publications across multiple clinical domains, including a machine learning study using the RACH service for early Alzheimer’s disease prediction published in the Journal of Prevention of Alzheimer’s Disease,^18^ and a study modeling COVID-19 disease outcomes published in Emerging Microbes & Infections.^19^ This survey represents the first systematic capture of research outcomes at CBMI, a capability not available under prior workflows.

Apart from local service-tier research, HABITAT’s first-hand collaborations have contributed to multi-site studies, including a long COVID cohort study in children published in The Lancet Infectious Diseases,^20^ where HABITAT’s unstructured data anonymization pipeline enabled MU’s participation.

## Discussion

HABITAT grew out of a challenge common across academic medical centers: the growing gap between researcher demand for clinical data and the capacity of traditional informatics infrastructure to keep pace.^1^ Beyond operational throughput, HABITAT has begun to demonstrate measurable research value. Annual internal data request volume grew substantially relative to the pre-HABITAT era, now handled through a unified intake that consolidated fulfillment previously distributed across nine tools and three teams. Fulfillment efficiency improved despite a more complex request mix, and HABITAT-supported work has produced peer-reviewed publications in high-impact venues across multiple clinical domains. Beyond local research, HABITAT supported 29 externally funded collaborative projects across multiple federal, state, and industry sponsors, serving as a participating or lead site and providing governed data and analytic support to each project. This sustained multi-agency engagement reflects HABITAT’s role not only in fulfilling discrete requests but in enabling and supporting funded research and multi-site collaboration over time. These early outcomes, captured through a tracking system that did not previously exist, indicate that governance-first infrastructure delivers research value rather than imposing only administrative overhead.

Three years of operating HABITAT produced lessons relevant beyond MU. Curated data requests can require anywhere from 10 hours to several weeks of honest broker effort depending on phenotype complexity, IRB requirements, and data source integration, which is exactly why a sustainable cost model is needed from the start.^21^ Without one, honest broker teams take on escalating effort without cost recovery, creating risk of staff strain and service degradation.^22^ This challenge is compounded by the growing adoption of AI and big data analytics, which introduce substantial compute, storage, and infrastructure costs that are difficult to forecast and rarely accounted for in grant budgets. Many investigators do not account for informatics effort and technology costs when writing grant proposals, leaving informatics teams to cover costs that were never budgeted, a challenge NIH’s Data Management and Sharing Policy begins to address by explicitly permitting budgeting for informatics services.^23^ Working with informatics teams during grant development rather than after funding is secured matters for both research success and service sustainability.

Several peer institutions have independently developed components of this model such as ARCH at Weill Cornell Medicine,^24^ and Rush University’s Learning Healthcare System Data Commons.^25^ HABITAT’s contribution lies not in any single component but in integrating all three tiers under a single governed infrastructure where Five Safes-aligned^16^ controls operate as part of provisioning rather than as a separate downstream approval step. Our ecosystem is extended to national network research through PCORnet^4^ and GPC^3^, supporting use cases from coursework and dissertation work to funded research and multi-site collaborative projects. The 3.2% rejection rate and decrease in Tier 3 cancellations following clearer scope definition at intake both suggest that governance up front reduces requests that stall or get cancelled midway,^26^ though this model does add coordination burden and continued reliance on honest broker staff, a tradeoff CBMI continues to weigh as request volume grows. The consolidation of nine fragmented tools into a single governed pipeline also improved reproducibility, with each request traceable by Service ID and eligible cohorts retained within dedicated schemas, supporting traceability and auditability that the prior multi-tool process could not guarantee.

HABITAT’s data lakehouse architecture supported both local and national research beyond standard CDM workflows. As a participating site in the NIH RECOVER study, CBMI deployed its unstructured data anonymization pipeline on clinical notes before submission to the coordinating site.^20^ This pipeline was further developed for the ARPA-H-funded CADIF project, which also used non-standardized structured data elements from the HABITAT data model outside CDM scope. The ARPA-H-funded Cumulus project uses LLM-based workflows with health information exchange data, with HABITAT supporting local participation. The CDC-funded ALS4M study uses LLM-based pipelines on unstructured clinical data within HABITAT’s isolated compute environment. CBMI also piloted LLM-based concept extraction and clinical text exploration workflows internally within the same environment.

Several limitations encountered by CBMI that are worth noting are described in this section. Like many real-world data environments, HABITAT depends on source systems with incomplete documentation and variable use of coding standards.^27^ Some source elements remain unmapped to standard terminologies, and inconsistencies in upstream system documentation complicate standardization efforts, limiting the utility of certain data elements for network research. Security and privacy safeguards embedded within the provisioning workflow can also introduce additional review and coordination steps, occasionally extending request timelines when projects require detailed access assessments or cross-system approvals. Finally, the cost recovery model for RACH creates barriers for unfunded researchers, a challenge reported by other institutions balancing long-term service costs with equitable researcher access.^28^

Looking ahead, priorities include standardization of free text fields, broader ATLAS^12^ deployment, evaluation of cost recovery impacts on utilization, automation of intake steps where appropriate, and mapping of remaining data elements to standard coding systems to improve interoperability and support multi-site research. Our future work also includes expanding LLM support, analytical federations and rapid feedback cycles to improve data quality.

## Conclusion

HABITAT demonstrates that a governance-first clinical research data infrastructure can support diverse investigator needs within a unified, tiered ecosystem, reducing reliance on fragmented, project-specific data solutions. By embedding structured intake, IRB alignment, honest broker workflows, and Five Safes-aligned controls, it treats patient privacy and data security as part of data access rather than external constraints. Over three years of operation, this approach has supported both self-service and curated data access, local and multi-site collaborative research, and emerging use cases including projects leveraging large language models, showing that advanced analytics and network science can operate within governed environments. Several elements of HABITAT, including the three-tier service architecture, governance workflow, REDCap intake system, and AWS/Snowflake infrastructure built on open standards (PCORnet CDM, OMOP, LOINC), are designed for broader applicability and could be adapted by other academic medical centers seeking to modernize their research data infrastructure. As AI and big data analytics place increasing computational and financial demands on research infrastructure, governance-first models with transparent cost recovery frameworks will be critical to sustaining research data services over time.

## Data Availability

All data produced in the present study are available upon reasonable request to the authors.

## Acknowledgements

The development of HABITAT was supported by the MU School of Medicine Dean’s Office, PCORI Program Award RI-MISSOURI-01-PS8, ARPA-H (D24AC00413-00), and AWS Global Health Equity Award. We are grateful to Dr. Russ Waitman and Dr. Abu Saleh Mosa for their leadership and contributions during the early development of HABITAT at MU. We thank the MU compliance, security, and IRB teams for working closely with us to ensure the governance frameworks. We also acknowledge the Tiger Institute for their ongoing partnership in providing access to raw EHR source data and Snowflake for infrastructure support.

